# Perioperative albumin versus other fluids to prevent cardiac surgery associated kidney injury: a protocol for a systematic review and meta-analysis of randomised trials

**DOI:** 10.1101/2024.09.05.24313089

**Authors:** Phoebe Darlison, Alastair Brown, Ary Serpa Neto, Adrian Pakavakis, Mayurathan Balachandran, Yahya Shehabi

## Abstract

**Background:** Acute kidney injury is a common complication following cardiac surgery. Albumin infusions have been proposed as an intervention that reduce the risk of this complication, but existing data have shown heterogenous results. The recent completion of two randomised controlled trial of Albumin infusions in cardiac surgical patients provides the opportunity to conduct a systematic review and meta-analysis to improve the precision of the estimated treatment effect of Albumin infusions in cardiac surgery.

**Methods:** We will conduct a systematic review of randomised controlled trials that have evaluated the use of peri-operative Albumin infusions compared to comparator fluids in patient undergoing on-pump cardiac surgery. We will conduct a search of MEDLINE, EMBASE, CINAHL and Cochrane Central from inception to 22nd August. The review will be conducted and reported in accordance with the PRISMA 2020 statement. We will use a bayesian framework to estimate the treatment effect of albumin to prevent acute kidney injury defined according to the KDIGO criteria.

**Results:** This systematic review has been prospectively registered on PROSPERO (CRD42024580170) and the formal search was conducted on 23^rd^ August 2024. Title and abstract screening has commenced with data extraction to commence following the submission of the protocol.

**Conclusion:** This systematic review will provide an updated systematic review and meta-analysis to inform clinicians about the role Albumin infusion in cardiac surgery.

## Background

Cardiac surgery associated Acute Kidney Injury (CSA-AKI) is a common complication of cardiac surgery and affects 20 -40% of patients undergoing surgery^1-3^. CSA-AKI has been shown to be associated with adverse patient-centred outcomes including increased risk of mortality and long term renal impairment^2,4-6^. The administration of intravenous solutions to optimise fluid balance is recognised as a key method of preventing acute kidney injury with both hypovolaemia and hypervolaemia associated with increased risk^1-3,7^. Until recently no interventions had been shown to reduce the risk of CSA-AKI. However the PROTECTION study demonstrated an intravenous amino acid infusion reduced the risk of CSA-AKI supporting the potential role of protein containing solutions in improving renal outcomes after cardiac surgery^8^.

Human albumin solutions are protein containing fluids that are commonly administered to patients both during and after cardiac surgery. Albumin solutions have been shown to potentially optimise intravascular volume through the maintenance of colloid oncotic pressure and reduce the accumulation of positive fluid balance^9^. Albumin solutions have also been suggested to potentially improve renal blood flow autoregulation, reduce oxidative stress, and stabilise the endothelial glycocalyx^10-14^. Despite these proposed benefits a recently published systematic review highlighted the equivocal results of albumin solutions on renal function in the cardiac surgery and vascular population^15^.

Similarly, the International Collaboration for Transfusion Medicine Guidelines (ICTMG) have made a weak recommendation against the use of Albumin to prevent acute kidney injury in major surgery^16^. However both the guideline and the review were limited by the heterogenous populations, small sample sizes and the low incidence of acute kidney injury in the included studies.

Since these publications, the 20% Human Albumin Solution Bolus Fluid Administration Therapy After Cardiac Surgery (HAS FLAIR) II^17^ and 20% Albumin and Acute Kidney Injury (ALBICS-AKI) trials^18^, have been completed, and provide data for over 1000 additional patients. Therefore, we plan to perform this updated systematic review and Bayesian meta-analysis to assess whether Albumin administration during and after cardiac surgery is associated with a reduced risk of CSI-AKI and other clinical outcomes.

## Objectives

The objective of this systematic review is to evaluate the impact of intraoperative and postoperative albumin fluid therapy when compared to other fluid regimes on the risk of acute kidney injury and other clinical outcomes in patients who have undergone cardiac surgery on cardiopulmonary bypass.

## Methods

### Selection of studies

We will include randomised controlled trials comparing any albumin-containing fluid therapy with any comparator fluid regime given either intraoperatively or postoperatively in adult patients (^3^18 years old) undergoing on-bypass cardiac surgery. Studies will be included irrespective of publication status or publication date. This will include unpublished studies and full-text publications. Conference abstracts will be excluded. A detailed description of the definitions of the types of studies, participants and interventions that will be used is included in appendix 2. We will include studies which include at least one of the primary outcome or secondary outcome measures.

### Outcome Measures

The primary outcome measure will be the incidence of acute kidney injury within the hospital admission. Definitions used by trial authors will be unified using the KDIGO criteria (see appendix 2 for details)^19^. The secondary outcomes we will collect will be all-cause mortality at longest follow-up, the proportion of patients requiring renal replacement therapy postoperatively, the duration of invasive ventilation postoperatively, ICU and hospital length of stay, and the duration of inotrope and/or vasopressor therapy.

### Identification of Studies

We will perform a search of the MEDLINE, CINAHL, EMBASE and the Central Registrar of Controlled Trials (CENTRAL) from inception to the date of the search as described in the Cochrane Handbook of Systematic Reviews of Interventions Chapter 4^20^. There will be no language, publication year or publication status restrictions. Our search strategy has been developed in conjunction with a research librarian and subject matter experts. A draft search strategy for MEDLINE and Embase is included in appendix 1. A full search strategy will be submitted with the final publication. We will check bibliographic references and citations of relevant studies and reviews for further references to trials which may be eligible for inclusion. We will also search the Australian New Zealand Clinical Trials Registry (ANZCTR), ClinicalTrials.gov, the World Health Organization International Clinical Trial Registry Platform, and the ISRCTN registry for unpublished and ongoing studies^21-24^. We will contact trial authors when necessary for further information.

### Data collection and analysis

#### Selection of studies

Screening and data extraction will be completed and documented according to the PRISMA 2020 statement using the Covidence systematic review tool^25,26^. Duplicate extract and non-randomised controlled trials will be automatically excluded using Covidence. Titles and abstracts of all remaining records retrieved during the search process will be independently reviewed by two of the review authors to identify potentially eligible studies. Full-text publications or study reports will then be retrieved and screened independently by two of the review authors to identify the studies which meet the inclusion criteria. Any disagreements during the screening process will be resolved through discussion between reviewers until a resolution is achieved, and if necessary, by involvement of the senior author (YS). We will identify and exclude duplicate publications. We will identify multiple reports or publications of the same trial, and collate reports so that each study, rather than each report are reviewed ensuring the data are not duplicated.

#### Data extraction and management

Data extraction and management will be performed using the Covidence ‘Extraction 1’ data system^25^. Data from each included study will be extracted in duplicate by two independent reviewers using a pre-defined data extraction template based on the review inclusion criteria and recommendations in Chapter 5 of the Cochrane Handbook for Systematic Reviews of Interventions^27^. The data extraction template will be piloted by at least two reviewers prior to use. Disagreements in data extraction will be resolved in discussion between the two reviewers. If required involvement of a third reviewer will be initiated to mediate and come to a conclusion regarding the disagreement. Trial investigators of included studies will be contacted via email to request details or clarification regarding any missing data that are identified during the data extraction process. The investigators of the included trials will be contacted a maximum of two times, if no reply is returned within a reasonable timeframe, the data will be reported as missing for the meta-analysis.

#### Assessment of risk of bias in included studies

Risk of bias for each study will be assessed using the Cochrane Risk of Bias tool for randomized trials (RoB 2)^28,29^. Risk of bias will be assessed by two independent reviewers in duplicate in the five domains included in the RoB2 tool. The judgements of the two reviewers will then be compared, and disagreements between judgements will be resolved with discussion between the reviewers.

#### Measures of treatment effect

Effect sizes will be presented as risk ratios (RRs) for binary outcomes and mean differences for continuous outcomes. We will evaluate the treatment effect using the intention to treat populations.

#### Unit of analysis

The unit of randomisation in this review is anticipated to be the trial level.

#### Assessment of heterogeneity

Quantitative heterogeneity will be assessed with the posterior estimates of the heterogeneity parameter (τ) with its 95% CrI. Subgroup heterogeneity will be assessed by including an interaction term in the analysis to obtain an estimate and 95% CrI for the ratio of RRs (RRRs) from the posterior distribution of the interaction estimate.

#### Assessment of reporting biases

Where at least 10 studies are available for meta-analysis, we will use funnel plots to assess for small study effects to evaluate whether publication bias has affected the review as a whole. We will review the included studies to assess whether studies may be duplicate publications of the same participant cohort and contact authors for clarification if this is unclear.

#### Data synthesis

A bayesian framework will be used as the primary statistical approach.

Pooled estimates of effect sizes as risk ratios (RRs) for binary outcomes, and mean differences for continuous outcomes will be reported. Continuous variables presented in formats not readily amenable to pooling will be converted to mean and SD with the method described elsewhere^30^. Along with the pooled estimates of effect sizes, 95% credible intervals (CrIs) calculated using the shortest interval method, which for unimodal posteriors is equivalent to the highest posterior density region method will be presented. For all analyses, Bayes factors will be based on marginal likelihoods. Trials with zero events (if any) will be included in the final model and an effect estimate calculated accordingly.

All analyses will be performed considering a minimally informative (unit information prior for the log-RR) distribution for the effect prior, and a weakly informative half-normal prior distribution with scale 0·5 for the heterogeneity prior. Priors were selected on the basis of previous recommendations^31,32^. The treatment effect prior probability distribution will be defined by setting an optimistic, a pessimistic, and a minimally informative prior belief for the treatment effect^33^. The optimistic and pessimistic priors will be used only in sensitivity analyses and the main analysis will use the minimally informative prior. The strength of these prior beliefs (the variance setting to establish the shape of the distribution) will be set as moderate for the optimistic and minimally informative priors and weak for the pessimistic prior. In other words, the priors will be set so that we cannot rule out an eventual benefit but can mostly rule out large effect sizes for the intervention and acknowledge a non-negligible chance of the intervention being harmful. In mathematical form, the minimally informative prior will be normally distributed and centred at the absence of effect [OR = 1; log(OR) = 0] with a standard deviation (SD) of 0.355, such that 0.95 of the probability falls in the range of 0.5–2. The pessimistic and optimistic priors will be informed by the range of effect size estimates from previous studies suggesting the effect of albumin ranging from a reduction of 10% in the risk of AKI to an increase of 3%^34,35^ (OR = 0.90 for the optimistic prior and OR = 1.03 for the pessimistic prior, Figure 1). The optimistic prior SD will be defined to retain a 0.15 probability of harm [Pr(OR > 1)] (SD = 0.10), and the pessimistic prior will be defined to retain a 0.30 probability of harm [Pr(OR < 1)] (SD = 0.06). All statistical analyses will be performed with R version 4.3.3 (R Foundation for Statistical Computing) using the bayesmeta package^36,37^.

**Figure 1:**
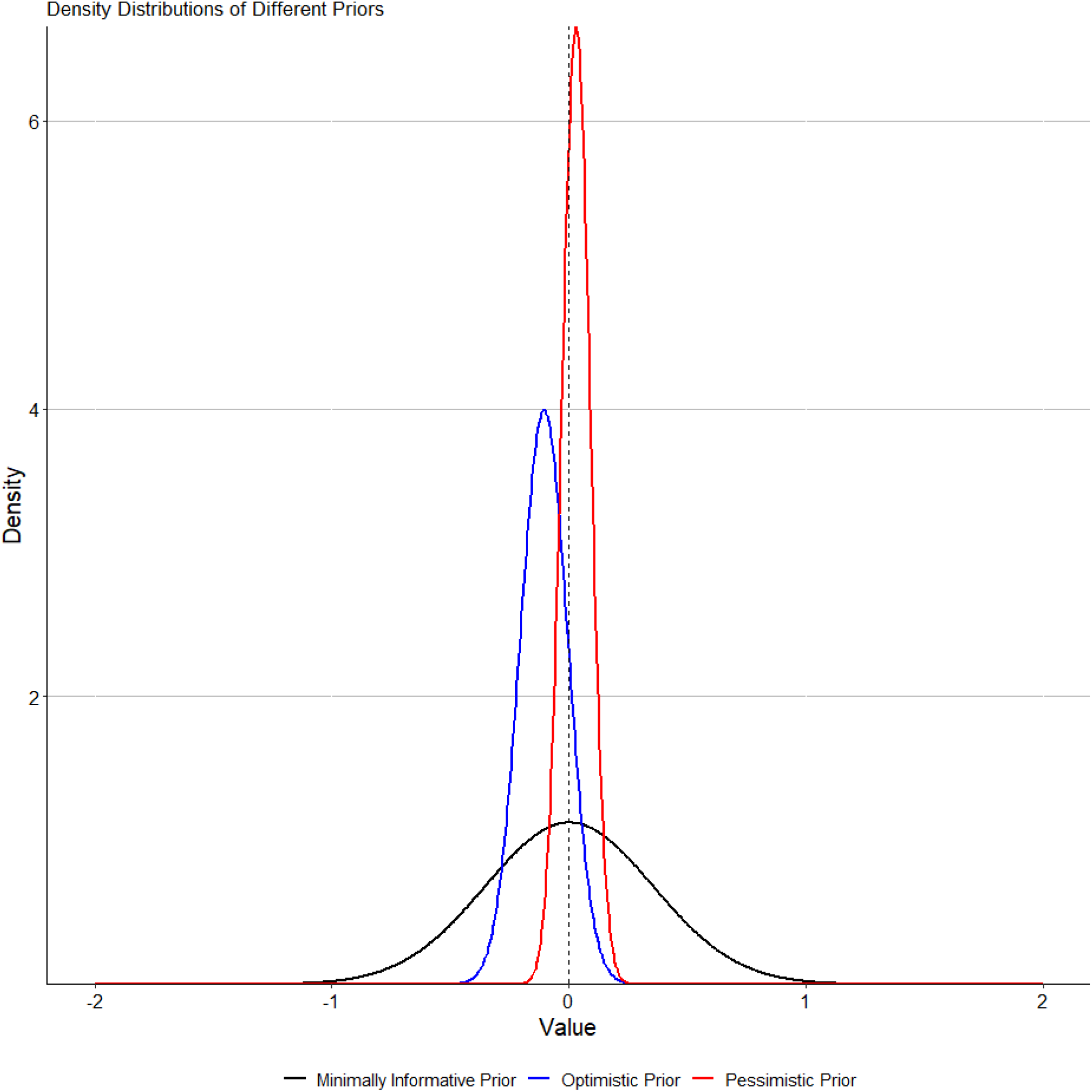
Density Distributions of Priors for effect estimate analyses

#### Subgroup analysis and assessment of heterogeneity

We will perform the following subgroup analyses to examine for differential effects of:

- Different concentrations of albumin (4-5% albumin and 20-25% albumin) and different comparator fluids (crystalloid and colloid fluids)
  - 4-5% albumin versus comparator fluids
  - 20-25% albumin versus comparator fluids
- Timing of Albumin therapy
  - Intraoperatively only (including for priming)
  - Both intraoperative and postoperative
- Different risk populations
  - Populations with eGFR <60mL/min/1.73m2 preoperatively
  - Combined procedures

#### Sensitivity analysis

Sensitivity analyses examining treatment effects using different priors of effect and heterogeneity parameters will be conducted. The sensitivity analysis will involve the following reanalyses:

- Excluding studies with high risk of bias
- Using informative priors as described above

#### Summary of findings table and GRADE

We will assess certainty of evidence using the GRADE criteria (gdt.gradepro.org) in the five GRADE considerations (risk of bias, consistency of effect, imprecision, indirectness, publication bias)^38^. We will create ‘summary of findings’ tables for the following key outcomes and comparisons which are most relevant to stakeholders. Outcomes:

- Acute kidney injury within hospital stay
- Renal replacement therapy
- All-cause mortality at longest follow-up
- ICU length of stay
- Duration of vasopressor therapy

Comparisons:

- 4-5% albumin versus comparator fluids
- 20-25% albumin versus comparator fluids

## Data Availability

All data produced in the present work are contained in the manuscript

## Acknowledgements

Thank you to Anna Lovang and the St Vincent’s Hospital Melbourne Library for assistance and guidance in developing search strategies and assisting with advanced research support.

