## Supplemental File for "Perioperative albumin versus other fluids to prevent cardiac surgery associated kidney injury: a protocol for a systematic review and meta-analysis of randomised trials"

### Appendix 1: Search Strategy Examples

#### MEDLINE Search Strategy Example

| | | |
|---|---|---|
| **Database Search:** Ovid MEDLINE(R) ALL<br>**Search Date:** 19/08/2024<br>**Dates:** 1946 to August 21, 2024<br>**Language:** Any | | |
| Search No. | Search Terms | Results |
| 1 | exp Fluid Therapy/ or Fluid Therap*.mp. | 24903 |
| 2 | Fluid Regime*.mp. | 411 |
| 3 | Fluid Bolus*.mp. | 1030 |
| 4 | Intravenous*.mp. or exp Infusions, Intravenous/ | 496617 |
| 5 | Infusion*.mp. | 327361 |
| 6 | Colloid*.mp. or exp Colloids/ | 222627 |
| 7 | exp Crystalloid Solutions/ or Crystalloid*.mp. | 11136 |
| 8 | Crystalloid Solution*.mp. | 2792 |
| 9 | Balanced Salt Solution*.mp. | 3450 |
| 10 | Balanced Crystalloid*.mp. | 378 |
| 11 | exp Isotonic Solutions/ or Isotonic Solution*.mp. | 11885 |
| 12 | exp Solutions/ | 138523 |
| 13 | Solution*.mp. | 959424 |

| | | |
|---|---|---|
| 14 | exp Plasma Substitutes/ or Plasma Substitute*.mp. | 40688 |
| 15 | Hypertonic Solution*.mp. | 6789 |
| 16 | Hypotonic Solution*.mp. | 3873 |
| 17 | Volume Expansion*.mp. | 8362 |
| 18 | exp Sodium Chloride/ or Sodium Chloride*.mp. | 87140 |
| 19 | Saline*.mp. or exp Saline Solution/ | 209575 |
| 20 | Normal Saline*.mp. | 26965 |
| 21 | Hartmann Solution*.mp. | 20 |
| 22 | Hartman*.mp. | 5964 |
| 23 | exp Sodium Lactate/ or Compound* Sodium Lactate.mp. | 422 |
| 24 | Lactate Sodium*.mp. | 121 |
| 25 | Plasmalyte*.mp. | 201 |
| 26 | exp Albumins/ | 200914 |
| 27 | Albumin*.mp. | 237009 |
| 28 | exp Serum Albumin/ or Serum Albumin*.mp. | 129674 |
| 29 | Human Albumin*.mp. | 3438 |
| 30 | Human Serum Albumin*.mp. or exp Serum Albumin, Human/ | 23371 |
| 31 | Intravenous Albumin*.mp. | 294 |
| 32 | exp Starch/ or Starch*.mp. | 91184 |

| | | |
|---|---|---|
| 33 | exp Hydroxyethyl Starch Derivatives/ or Hydroxyethyl*.mp. | 21037 |
| 34 | Hydroxyethyl*.mp. | 21037 |
| 35 | Hetastarch*.mp. | 514 |
| 36 | exp Ringer's Solution/ or Ringer*.mp. | 15310 |
| 37 | exp Ringer's Lactate/ or Ringer* Lactate.mp. | 3054 |
| 38 | exp Gelatin/ or Gelatin*.mp. | 59953 |
| 39 | exp Succinates/ or Gelatin Succinate*.mp. | 25181 |
| 40 | Succinate*.mp. | 47486 |
| 41 | exp Polygeline/ or Polygeline*.mp. | 375 |
| 42 | Glucose*.mp. or exp Glucose/ | 710308 |
| 43 | Dextrose*.mp. | 15699 |
| 44 | exp Dextrans/ | 34725 |
| 45 | Dextran*.mp. | 58250 |
| 46 | exp Cardiac Surgical Procedures/ or Cardiac Surg*.mp. | 278128 |
| 47 | exp Cardiovascular Surgical Procedures/ or Cardiovascular Surg*.mp. | 469742 |
| 48 | Cardiothoracic Surg*.mp. | 5415 |

| | | |
|---|---|---|
| 49 | exp Thoracic Surgical Procedures/ or Thoracic Surg*.mp. | 399129 |
| 50 | Heart Surg*.mp. | 22572 |
| 51 | Open Heart Surg*.mp. | 10055 |
| 52 | exp Extracorporeal Circulation/ or Extracorporeal Circ*.mp. | 99133 |
| 53 | exp Cardiopulmonary Bypass/ or Cardiopulmonary Bypass*.mp. | 44172 |
| 54 | exp Myocardial Revascularization/ or Myocardial Revasc*.mp. | 100222 |
| 55 | Heart Muscle Revasc*.mp. | 0 |
| 56 | exp Coronary Artery Bypass/ or Coronary Artery Bypass*.mp. | 74575 |
| 57 | Coronary Artery Surg*.mp. | 2298 |
| 58 | Aorta Surg*.mp. | 826 |
| 59 | Aortic Surg*.mp. | 4882 |
| 60 | Ascending Aorta Surg*.mp. | 73 |
| 61 | Heart Valve Surg*.mp. | 887 |
| 62 | Cardiac Valve Surg*.mp. | 443 |
| 63 | exp Intraoperative Care/ or exp Intraoperative Period/ or Intraoperative*.mp. | 247462 |

| | | |
|---|---|---|
| 64 | Perioperative*.mp. or exp Perioperative Care/ or exp Perioperative Period/ | 364525 |
| 65 | Peroperative*.mp. | 4614 |
| 66 | exp Postoperative Period/ or exp Postoperative Care/ or Postoperative*.mp. | 1044695 |
| 67 | exp Postoperative Complications/ or Postoperative Complication*.mp. | 682911 |
| 68 | exp Postoperative Hemorrhage/ | 13649 |
| 69 | Perioperative Complication*.mp. | 13981 |
| 70 | 1 or 2 or 3 or 4 or 5 or 6 or 7 or 8 or 9 or 10 or 11 or 12 or 13 or 14 or 15 or 16 or 17 or 18 or 19 or 20 or 21 or 22 or 23 or 24 or 25 or 32 or 33 or 34 or 35 or 36 or 37 or 38 or 39 or 40 or 41 or 42 or 43 or 44 or 45 | 2881940 |
| 71 | 26 or 27 or 28 or 29 or 30 or 31 | 326331 |
| 72 | 70 and 71 | 69693 |
| 73 | 46 or 47 or 48 or 49 or 50 or 51 or 52 or 53 or 54 or 55 or 56 or 57 or 58 or 59 or 60 or 61 or 62 | 733113 |
| 74 | 63 or 64 or 65 or 66 or 67 or 68 or 69 | 1411513 |

| | | |
|---|---|---|
| 75 | 73 or 74 | 1940440 |
| 76 | 72 and 75 | 4214 |

#### EMBASE Search Strategy Example

**Database Search:** Ovid EMBASE

**Search Date:** 19/08/2024

**Dates:** 1974 to August 16, 2024

**Language:** Any

| Search No. | Search Terms | Results |
|---|---|---|
| 1 | exp fluid therapy/ or fluid therap*.mp. | 119459 |
| 2 | infusion*.mp. or exp infusion fluid/ or exp infusion therapy/ | 501550 |
| 3 | intravenous*.mp. or exp intravenous drug administration/ | 1351794 |
| 4 | fluid regime*.mp. | 545 |
| 5 | fluid bolus*.mp. | 2183 |
| 6 | exp colloid/ or colloid*.mp. | 99188 |
| 7 | exp crystalloid/ or crystalloid*.mp. | 15870 |
| 8 | exp balanced salt solution/ or balanced salt solution*.mp. | 27151 |

| | | |
|---|---|---|
| 9 | exp isotonic solution/ or isotonic solution*.mp. | 5027 |
| 10 | exp "solution and solubility"/ | 306397 |
| 11 | solution*.mp. | 1094720 |
| 12 | hypertonic solution*.mp. | 5458 |
| 13 | hypotonic solution*.mp. | 2944 |
| 14 | exp plasma substitute/ or plasma substitute*.mp. | 76876 |
| 15 | volume expansion*.mp. | 9652 |
| 16 | exp sodium chloride/ or sodium chloride*.mp. | 249306 |
| 17 | saline*.mp. | 293786 |
| 18 | normal saline*.mp. | 42472 |
| 19 | exp Hartmann solution/ or hartmann solution*.mp. | 799 |
| 20 | hartman*.mp. | 9442 |
| 21 | lactate sodium*.mp. or exp lactate sodium/ | 1759 |
| 22 | compound* sodium lactate*.mp. | 51 |
| 23 | exp acetic acid plus gluconate sodium plus magnesium chloride plus potassium chloride plus sodium chloride/ or plasmalyte*.mp. | 1036 |
| 24 | exp albumin/ or albumin*.mp. | 372419 |

| | | |
|---|---|---|
| 25 | exp serum albumin/ or serum albumin*.mp. | 126187 |
| 26 | exp human albumin/ or human albumin*.mp. | 6015 |
| 27 | exp human serum albumin/ or human serum albumin*.mp. | 23099 |
| 28 | intravenous albumin*.mp. | 469 |
| 29 | starch*.mp. or exp starch/ | 69511 |
| 30 | exp hydroxyethyl starch/ or hydroxyethyl starch*.mp. | 4548 |
| 31 | hetastarch*.mp. or exp hetastarch/ | 7925 |
| 32 | hydroxyethyl*.mp. | 35360 |
| 33 | exp hydroxyethylcellulose/ or hydroxyethylcellulose*.mp. | 2147 |
| 34 | ringer solution*.mp. or exp Ringer solution/ | 5669 |
| 35 | exp Ringer lactate solution/ or ringer* lactate solution.mp. | 10177 |
| 36 | exp Ringer acetate/ or ringer acetate*.mp. | 632 |
| 37 | ringer*.mp. | 24355 |
| 38 | exp gelatin/ or gelatin*.mp. | 164819 |

| | | |
|---|---|---|
| 39 | exp gelatin succinate/ or gelatin succinate*.mp. | 939 |
| 40 | exp gelatin plus sodium chloride/ or gelatin plus sodium chloride*.mp. | 4 |
| 41 | exp polygeline/ or polygeline*.mp. | 1027 |
| 42 | exp glucose/ or glucose*.mp. or exp glucose infusion/ | 1078205 |
| 43 | exp glucose plus sodium chloride/ or glucose plus sodium chloride*.mp. | 75 |
| 44 | dextrose*.mp. | 20744 |
| 45 | exp dextran/ or dextran*.mp. | 72665 |
| 46 | exp heart surgery/ or heart surg*.mp. | 475542 |
| 47 | exp cardiovascular surgery/ or cardiovascular surg*.mp. | 913787 |
| 48 | cardiac surg*.mp. | 92503 |
| 49 | cardiothoracic surg*.mp. | 9484 |
| 50 | exp thorax surgery/ or thorax surg*.mp. | 725220 |
| 51 | exp open heart surgery/ or open heart surg*.mp. | 16901 |
| 52 | exp cardiopulmonary bypass/ or | 76284 |

| | | |
|---|---|---|
| | cardiopulmonary bypass*.mp. | |
| 53 | exp extracorporeal circulation/ or extracorporeal circ*.mp. | 87273 |
| 54 | exp heart muscle revascularization/ or heart muscle revasc*.mp. | 40100 |
| 55 | myocardial revasc*.mp. | 7693 |
| 56 | exp coronary artery bypass graft/ or coronary artery bypass graft*.mp. | 105764 |
| 57 | exp coronary artery surgery/ or coronary artery surg*.mp. | 157499 |
| 58 | exp aortic surgery/ or aortic surg*.mp. | 48492 |
| 59 | exp aorta surgery/ or aorta surg*.mp. | 51389 |
| 60 | exp ascending aorta surgery/ or ascending aorta surg*.mp. | 2494 |
| 61 | exp heart valve surgery/ or heart valve surg*.mp. | 138293 |
| 62 | intraoperative period.mp. or exp intraoperative period/ | 278224 |
| 63 | intraoperative*.mp. | 285696 |
| 64 | perioperative*.mp. or exp perioperative complication/ or exp | 1901126 |

| | | |
|---|---|---|
| | perioperative care/ or exp perioperative period/ | |
| 65 | peroperative care.mp. or exp peroperative care/ | 15694 |
| 66 | exp peroperative complication/ or peroperative*.mp. | 89998 |
| 67 | postoperative*.mp. or exp postoperative care/ or exp postoperative period/ or exp postoperative monitoring/ | 1625088 |
| 68 | exp postoperative complication/ or postoperative complication*.mp. | 913168 |
| 69 | exp postoperative hemorrhage/ or postoperative hemorrhage*.mp. | 50896 |
| 70 | 1 or 2 or 3 or 4 or 5 or 6 or 7 or 8 or 9 or 10 or 11 or 12 or 13 or 14 or 15 or 16 or 17 or 18 or 19 or 20 or 21 or 22 or 23 or 29 or 30 or 31 or 32 or 33 or 34 or 35 or 36 or 37 or 38 or 39 or 40 or 41 or 42 or 43 or 44 or 45 | 4470240 |
| 71 | 24 or 25 or 26 or 27 or 28 | 372419 |
| 72 | 70 and 71 | 114725 |
| 73 | 46 or 47 or 48 or 49 or 50 or 51 or 52 or 53 or 54 or | 1205500 |

| | | |
|---|---|---|
| | 55 or 56 or 57 or 58 or 59 or 60 or 61 | |
| 74 | 62 or 63 or 64 or 65 or 66 or 67 or 68 or 69 | 2346878 |
| 75 | 73 or 74 | 3173728 |
| 76 | 72 and 75 | 10912 |

#### Appendix 2: Definitions

#### Experimental interventions

We will include studies where the intervention is perioperative intravenous fluid therapy with albumin-containing fluid.

Perioperative fluid therapy will be defined as intraoperative fluid administered as therapy (not including fluid used for priming of cardiopulmonary bypass only), or postoperative fluid therapy. Any dose or duration of intraoperative or postoperative fluid therapy will be eligible ensuring the intervention and comparator fluid type is eligible.

Intravenous albumin-containing fluid defined as any of the following preparations: 4% Albumin, 5% Albumin, 20% Albumin, or 25% Albumin.

#### Comparator interventions

We will include studies with perioperative fluid therapy with any comparator fluid regime that is different to the intervention fluid.

Perioperative fluid therapy will be defined as intraoperative fluid administered as therapy (not including fluid used for priming of cardiopulmonary bypass only), or postoperative fluid therapy.

#### Strategy for studies with a subset of eligible interventions

In studies where the intervention and/or comparator fluid is used for cardiopulmonary bypass priming and perioperative fluid therapy:

These studies will be included in the meta-analysis

A sensitivity analysis will be performed excluding the studies which have included intervention and/or comparator fluids used for cardiopulmonary bypass priming and perioperative fluid therapy

#### Types of outcome measures

##### Primary outcome measures

The primary outcome measure will be:

- Acute kidney injury within the hospital admission

- Definitions used by trial authors will be unified using the KDIGO criteria as described below[19]:
  - KDIGO stage 1 AKI corresponds with:
    - AKIN criteria stage 1
    - RIFLE criteria Risk category
  - KDIGO stage 2 AKI corresponds with:
    - AKIN criteria stage 2
    - RIFLE criteria Injury category
  - KDIGO stage 3 AKI corresponds with:
    - AKIN criteria stage 3
    - RIFLE criteria Failure, Loss, and End Stage categories
- Differences in classifications between trials will be reported

In studies where the intervention meets inclusion criteria, but renal outcomes are not reported, the study authors will be contacted for any renal data available.

##### Secondary outcome measures

We will include studies which include either the primary outcome measure or at least one of the following secondary outcome measures:

- All-cause mortality at longest follow-up
- Proportion of patients requiring renal replacement therapy postoperatively
- Duration of invasive ventilation postoperatively
- Duration of ICU admission postoperatively
- Duration of hospital admission postoperatively
- Duration of inotrope and/or vasopressor medication postoperatively
